# Estimating healthcare cost savings from greenspace in Washington

**DOI:** 10.64898/2026.10.21.26366190

**Authors:** Jingjing Tao, Sergey Rabotyagov, Matthew H. E. M. Browning, Gregory Bratman, Spencer A. Wood, Joshua Lawler

**Affiliations:** School of Environmental and Forest Sciences, University of Washington, Seattle, Washington, USA; Virtual Reality & Nature Lab, Department of Parks, Recreation & Tourism Management, College of Behavioral, Social and Health Sciences, Clemson University, Clemson, South Carolina, USA; Center for Nature and Health, University of Washington, Seattle, Washington, USA

**Keywords:** Nature and health, benefit transfer, greenspace, cost saving

## Abstract

Greenspace is increasingly recognized as public-health infrastructure, yet its potential contribution to healthcare cost savings remains poorly quantified beyond a relatively small number of studies. To contribute to these investigations, we estimated potential avoided healthcare expenditures associated with residential greenness in Washington using a benefit-transfer framework. We applied decile-specific relative healthcare-cost ratios from a Northern California cohort of approximately five million Kaiser Permanente members to population-weighted distributions of satellite-derived normalized difference vegetation index across Washington census block groups. Ratios were calibrated to Washington’s 2020 statewide per-capita healthcare expenditure, allowing estimates to reflect the state’s own cost baseline. Estimated annual per-capita costs declined from $6,472.75 in the least-green decile to $5,939.39 in the greenest, a difference of $533.35. Simulated interventions yielded estimated annual statewide savings of $45.6 million from raising the lowest decile to the next decile, $157.7 million from lifting the four lowest deciles to Decile five, and $56.9 million from urban-core greening in Washington’s five largest cities. These estimates are modeled associations, not causal forecasts, and depend on transferability between regional populations and on normalized difference vegetation index as an incomplete proxy for meaningful nature access. Nonetheless, the findings highlight increased and equitable greening efforts as a potentially consequential component of preventive health and infrastructure policy.

## Introduction

The connection between humans and the natural environment has long been recognized as fundamental and important. Human well-being is linked to nature through a complex architecture of ecosystem services that encompasses not only biophysical benefits, including climate stabilization, water purification, and flood protection, but also nonmaterial contributions to mental and physical health [1]. The concept of biophilia posits that human biological systems are evolutionarily attuned to natural stimuli, creating an adaptive dependence on intact ecosystems for optimal functioning [2,3]. Documented pathways through which greenspace affects health are multifaceted, spanning physiological, psychological, and behavioral domains. Physiologically, nature contact is associated with reduced allostatic load, a composite indicator of chronic stress burden, alongside improved neuroendocrine, metabolic, and immune biomarkers [4–7]. Psychologically, exposure to natural environments confers affective benefits, including decreased rumination, anxiety, and depressive symptoms [8,9]. At the behavioral level, natural environments function as affordances for increased physical activity, enhanced social cohesion, and reduced loneliness [10–12].

The urgency of understanding and measuring these linkages is underscored by contemporary patterns of global urbanization and climate change. While urban centers offer economic advantages, they are also associated with elevated burdens of some chronic non-communicable diseases, environmental stressors such as air pollution, noise, and the urban heat island effect, and mounting pressure on healthcare systems worldwide [5,13]. Climate change intensifies these urban environmental stressors unevenly: wildfire smoke, for instance, disproportionately degrades air quality in communities that already have limited access to parks and greenspace, compounding existing disparities in chronic disease burden [14]. Consequently, there is growing policy interest in using nature-based solutions and green infrastructure, such as parks, gardens, and street trees, as a proactive public health intervention to mitigate these effects and promote resilience. Scholars have accordingly advocated for an evidence-based reframing of greenspace as essential public health infrastructure capable of yielding substantial social returns on investment [15,16].

The existing literature has established robust associations between greenspace and a wide array of health outcomes. Research consistently links residential greenness to improved mental health, lower rates of cardiovascular disease, reduced risk of type 2 diabetes, and lower all-cause mortality [11,17,18]. Studies have examined diverse populations, including school-aged children, pregnant women, and veterans with post-traumatic stress disorder [10,19,20]. Methodological approaches have become sophisticated, drawing on high-resolution normalized difference vegetation index (NDVI) data, light detection and ranging (LiDAR)-based mapping, and longitudinal cohort tracking [7,17,21]. Causal links have been further strengthened by experimental and quasi-experimental studies, such as randomized controlled trials and difference-in-differences analyses of vacant lot greening, which have demonstrated significant reductions in feelings of depression and ambulatory heart rates within community participant samples [22,23].

Despite these advances, two compounding gaps remain in the literature. First, while the effect of nature on health is well recognized, few studies have translated these benefits into monetized healthcare cost savings, a gap consistent with a broader imbalance in the ecosystem services literature [24]. Valuation research remains concentrated on biophysical processes, such as carbon sequestration, with comparatively little economic research effort devoted to services tied to human health [25], even though credible dollar values are essential for these benefits to compete with other priorities in real decisions [26,27]. Recreation is one of the few nonmaterial benefits to be valued in monetary terms [28], although typically as willingness to pay and not nature’s health benefits specifically.

Second, even pioneering studies that have quantified healthcare cost savings from greenspace are geographically concentrated in a small number of settings: Australia [29,30] and the United States [31–34]. A recent systematic review synthesizing this evidence [24] likewise points to the narrowness of the geographic base underlying these findings. This leaves the transferability of healthcare cost savings from greenspace to other regions, including Washington, unexamined.

To help address these gaps, this study uses benefit transfer as a methodological bridge. We leverage the relative healthcare cost ratios across residential green cover deciles established in Northern California and apply them to Washington’s population-weighted greenness distribution. This approach requires the assumption that the fundamental relationship between green cover and health status, and its subsequent impact on healthcare use, is consistent across these two Western U.S. states. We acknowledge these assumptions and note that our study addresses them transparently through sensitivity analyses and context-specific demographic adjustments.

Washington provides a compelling location for this analysis due to its diverse geography and existing policy interest in the co-benefits of green infrastructure. Urban centers like Seattle have begun identifying opportunities to integrate nature into infrastructure planning to address health inequities and environmental stressors [35]. Applying benefit transfer here adds significant value by demonstrating how primary data from one region can be used to provide actionable economic evidence in another, thereby overcoming the high costs associated with primary longitudinal cost accounting. Our research objective is to quantify the potential avoided healthcare expenditures in Washington that could result from various levels of residential greenness exposure.

This study offers three specific contributions to the literature. Empirically, it provides the first greenspace healthcare cost savings estimates specifically for Washington, filling a critical regional data gap. Methodologically, it introduces a replicable benefit transfer framework that links satellite-derived green cover deciles to population-weighted cost estimates, offering a template for other states to conduct similar valuations. Finally, for policy, this study translates the multifaceted benefits of greenspace into the standardized economic language required for green infrastructure investment and public health budgeting. By doing so, we aim to frame natural spaces not merely as aesthetic amenities, but as public health infrastructure with potentially measurable economic benefits, supporting more equitable and sustainable urban development [15,36].

### Theoretical and mechanistic background

Foundational theoretical frameworks in environmental psychology include Stress Recovery Theory (SRT) [6] and Attention Restoration Theory (ART) [37]. SRT proposes that many types of nature contact elicit a precognitive, positive affective response that activates the parasympathetic nervous system, thereby facilitating physiological recovery from stress. ART posits that natural environments restore directed attention, a finite cognitive resource chronically depleted by urban demands, through the engagement of effortless soft fascination [3]. Importantly, the efficacy of restorative environments is not uniform across individuals – for example, biological responses are modulated by stress biology and life history, with early-life experiences shaping individual sensitivity to environmental stressors along an inverted-U dose-response curve [38].

Advances in environmental neuroscience and biomedicine have begun to isolate the specific mechanistic pathways through which nature impacts the body. One central candidate mechanism is the enhancement of immune functioning, particularly the activity of natural killer cells and the regulation of inflammatory cytokines [4,5]. These pathways may be activated by the inhalation of biogenic volatile organic compounds, such as forest terpenes, which have shown preliminary efficacy in reducing inflammatory markers [39]. Furthermore, short-term greenspace interventions have been shown to diversify the human gut and nasal microbiota, shifting composition toward beneficial taxa that promote immunoregulation [40,41]. Neuroimaging research complements these findings, demonstrating that current urban living is associated with increased amygdala activation during social stress, whereas exposure to urban greenspace can compensate for reduced prefrontal emotion-regulatory capacity in vulnerable individuals [9,42].

Beyond individual physiological responses, nature contact serves as a critical mediator of social health and behavioral choices. Longitudinal evidence indicates that time spent in natural environments can act as a scalable remedy for both social and emotional loneliness [43]. These environments foster social cohesion and perceived social support, which explain a significant portion of the association between greenspace and general health [10,12]. Moreover, the subjective experience of natural beauty has been shown to increase prosocial behaviors, including trust, generosity, and helping, mediated by the elicitation of positive emotions [44]. Such contact also influences long-term health behaviors; cross-national studies have identified a robust correlation between residential greenness and reduced prevalence of daily drinking and smoking [45].

The distribution of greenspace-related health benefits is also shaped by historical and structural determinants. Contemporary spatial inequities in urban greenspace can frequently be traced back to discriminatory mid-twentieth-century mortgage appraisal practices, including redlining, which produced lasting vegetation deficits in marginalized communities [46]. For residents of these neighborhoods, reduced greenspace access is compounded by disproportionate exposure to fine-particulate-matter air pollution, which is independently associated with elevated anxiety symptoms [47]. For Indigenous populations, the relationship between land access and health assumes additional dimensions of cultural identity and spiritual well-being and colonial dispossession and environmental degradation have been associated with profound and culturally specific forms of loss among Indigenous communities [48]. More broadly, disruptions to land and environment have also been linked to ecological grief and chronic climate-related psychological distress, which can affect any population facing environmental loss [49,50]. These structural realities point to an opportunity as much as a risk: greening interventions, when trauma-informed and culturally grounded, can help redress historical harms and expand capabilities, rather than reproduce displacement through processes of green gentrification [51,52].

## Materials and methods

In the fields of environmental and health economics, benefit transfer serves as a vital methodological bridge for policy evaluation. Benefit transfer is an established economic tool used to estimate the value of ecosystem services when primary data collection is either logistically infeasible or prohibitively expensive and involves the application of economic value estimates or functional relationships from a previously conducted source study to a new policy site where similar environmental or health attributes are under consideration [53,54]. This approach is particularly appropriate for our study because Washington lacks a large-scale, individual-level primary study that links residential greenspace exposure directly to longitudinal healthcare expenditure records and benefit transfer offers a practical means of generating policy-relevant estimates and interpretable relative cost indices for Washington despite this gap.

The methodological strength of this approach rests on two critical pillars of comparability: environmental context and health institutional structure. First, the source site (Northern California) and policy site (Washington) share comparable greenness and ecological characteristics. Both Northern California and Washington are situated within the broader Pacific Northwest and Cascadia bioregions and belong to the Pacific coastal temperate forest biome, sharing similar climate, vegetation, geology, and watershed processes and distribution. Furthermore, multiple U.S. Environmental Protection Agency Level III ecoregions, such as the Coast Range and the Eastern Cascades Slopes and Foothills, span both states, suggesting that the type of nature being experienced is biologically similar [55]. Second, the states possess comparable healthcare systems. They share similar institutional structures, including private insurance, Medicare and Medicaid programs, integrated healthcare delivery systems, and managed care organizations [56]. Notably, Kaiser Permanente, one of the nation’s largest integrated healthcare provider networks and the source of our primary estimates, operates extensively in both regions. According to Davis et al. [57]’s study, the provider serves approximately 4.4 million members in Northern California and 0.7 million members in Washington as of December 2020. This shared organizational infrastructure supports a degree of commonality in health system utilization patterns and provider structures across the two regions; moreover, Davis et al. [57] found that Kaiser Permanente’s regional membership is broadly comparable to the general population from which it is drawn on key sociodemographic characteristics, lending further support to the comparability of utilization patterns observed across regions.

The source for our transfer is the study conducted by Van Den Eeden et al. [33], which examined the relationship between residential greenness and healthcare costs among approximately 5 million Kaiser Permanente members in Northern California. This study is uniquely suited for transfer due to its large sample size and methodological transparency. The authors used a log-link gamma generalized estimating equation (GEE) to estimate the relative effect of greenness on healthcare costs, producing rate ratios (RRs) across ten deciles of the NDVI. These RRs represent the relative expected healthcare costs for individuals in greener environments compared to those in the least green decile (NDVI Decile 1). A key advantage of transferring these specific estimates is that the RRs were fully adjusted for a comprehensive suite of sociodemographic and environmental covariates, including age, sex, race/ethnicity, education, income, neighborhood deprivation, housing density, and air pollution. Because the source study estimated a common greenness effect after adjustment for sociodemographic covariates, and found these effects to be homogeneous across sociodemographic profiles without significant interaction terms, the relative effect of greenness on cost is assumed to be stable regardless of the specific demographic composition of our study site. The transferred cost ratios are multiplicative in the original log-link model, so the relative effect of greenness is invariant to the covariate values used in the original study. Thus, our transfer focuses on the greenness gradient alone without assuming that the Northern California healthcare cost production function applies to Washington. To preserve analytical validity, the following components of the source model are explicitly excluded from transfer: the regression intercept, Northern California baseline cost levels, Northern California covariate–cost relationships, and health system utilization patterns. These elements are population-and health-system-specific and cannot be reliably transported across geographic regions. Our analysis therefore relies exclusively on the decile-specific RRs and Washington’s own exposure distribution, avoiding ecological fallacies and false precision often associated with transferring absolute monetary values between different health systems.

To implement the transfer, we used high-resolution environmental and demographic data from Washington. The primary geographic unit of analysis is the census block group (CBG), which serves as an exposure assignment unit representing the typical greenness environment of their residents. CBGs are optimal for this framework because they offer reliable population counts and are sufficiently small to meaningfully reflect local variations in greenspace while aligning spatially with the resolution of satellite-derived vegetation data.

Greenness exposure was quantified using NDVI values derived from the moderate resolution imaging spectroradiometer (MODIS) 16-day composite reflectance products using 250-m, 500-m, and 1000-m buffers around CBG centroids. All available composites spanning the study period were acquired for tiles covering Washington, mosaicked to produce a spatially contiguous statewide raster, and pixel-wise annual temporal mean NDVI was computed across all composites to yield a time-averaged greenness exposure surface. Following the preprocessing protocol of our source study, we masked all values representing open water (NDVI < 0) to focus exclusively on terrestrial vegetation. NDVI values were used to assign each block group to one of ten population-weighted greenness deciles, the same decile structure used to estimate rate ratios in the source study. Washington appears greener than Northern California: over the past two decades, between 14% and 21% of the state’s land area has exceeded the greenness threshold applied in the source data, placing much of Washington’s population above the range that Northern California’s own decile structure was built to capture. Because no source-derived rate ratio exists beyond Northern California’s greenest decile, Washington block groups exceeding that maximum are assigned the source study’s top-decile ratio by default. This treatment produces a conservative estimate of total potential benefits only if the greenness–cost relationship continues to strengthen, rather than plateau or reverse, beyond the range observed in Northern California, an assumption we cannot test with the present data and therefore treat as a plausible interpretation rather than a demonstrated lower bound.

For each CBG *b*, let *N_b_* represent the resident population and *d* or *G_b_* ∈ {1,…,10} denotes the NDVI decile assigned to that block group, with the lowest-greenness decile serving as the reference category in the source study, and RR_Gb_or RR*_d_* the corresponding relative cost ratio for that decile. Let N = Σ_b_ N_b_ be the total Washington population, then the state population share in NDVI decile *d* is calculated as:

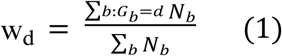

The resulting vector 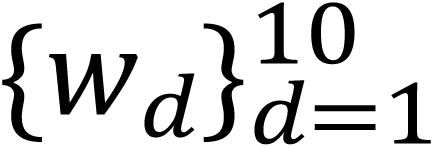 constitutes Washington’s population-weighted greenness distribution, constrained to satisfy 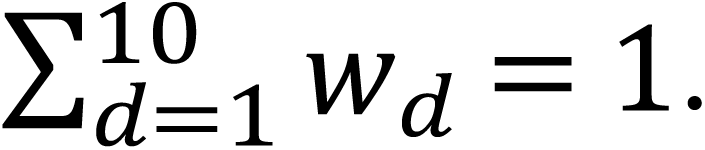 Because Washington’s data provide only the statewide average per-capita healthcare cost, denoted by 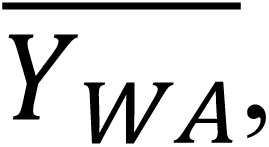 and not the absolute cost in the reference greenness decile, the transferred ratios must be normalized so that the population-weighted average of the predicted decile-specific costs equals the observed statewide mean. Specifically, let the expected per-capita healthcare cost in decile *d* be written as:

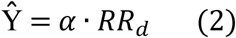

The expected per-capita healthcare cost in Washington is obtained by scaling the state’s baseline cost by the population-weighted average of the transferred relative ratios:

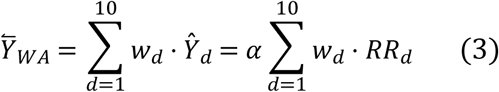

where *w_d_* represents the population-weighted exposure distribution across NDVI deciles in Washington. These weights form the basis for all state-level relative cost calculations. the statewide relative cost index for Washington is defined as:

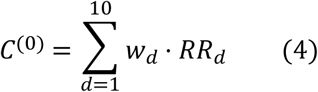

which is a dimensionless index representing expected per-capita healthcare costs in Washington relative to a hypothetical population uniformly assigned to the least-green decile in the Northern California reference sample. Rearranging Equation (3), the scaling constant *α*, which equals the expected per-capita cost in the reference decile (Decile 1, where *RR* = 1), is:

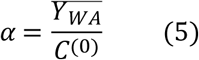

To quantify the potential healthcare cost savings attributable to greenspace investment, we evaluated three policy-relevant greening scenarios. For each scenario, a counterfactual population-weighted distribution 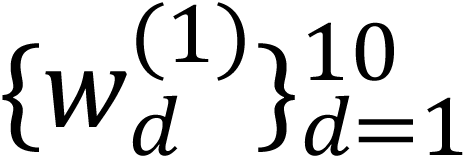 was constructed by reassigning block groups from lower to higher NDVI deciles according to the scenario specification. The counterfactual statewide cost index is:

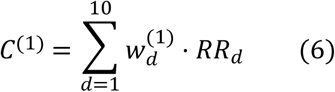

The percentage change in expected per-capita healthcare costs attributable to the greening scenario is:

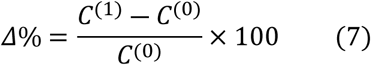

The approximate per-capita annual healthcare cost savings are then:

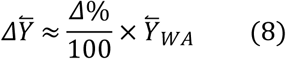

Aggregate statewide annual savings were obtained by multiplying the per-capita estimate by Washington’s total resident population *N_WA_*:

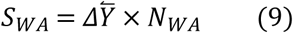

We examined three ecologically and policy-relevant scenarios using R and ArcGIS Pro to calculate simulation results. Scenario 1 (Bottom-Decile Lift) reallocates all CBGs currently in NDVI decile 1 to decile 2, simulating a targeted intervention in the least-green areas. Scenario 2 (Lower-Quintile Uplift) shifts all CBGs below the fifth NDVI decile to decile 5, representing a broader landscape greening intervention across the lowest four deciles of the greenness distribution. Scenario 3 (Urban Core Greening) targets CBGs located in the urban cores of Washington’s five largest cities by 2020 U.S. Census population: Seattle, Spokane, Tacoma, Vancouver, and Bellevue, applying the Scenario 2 reassignment rule to block groups in deciles 1–4 within these cities to simulate city-level green-infrastructure programs.

## Results and Discussion

Fig 1 presents the statewide NDVI distribution for Washington, divided into ten population-weighted deciles using 2020 CBG data. The distributional profile shows that compared to Northern California, the geographic context from which the source rate ratios were obtained, Washington’s residential landscape is greener. The upper half of the NDVI spectrum is occupied by a disproportionately high percentage of Washington’s census block groups and their inhabitants, which is indicative of the state’s temperate maritime climate, substantial coniferous forest cover, and well-preserved riparian corridors that span its eastern plateau and western lowlands.

**Fig 1.**
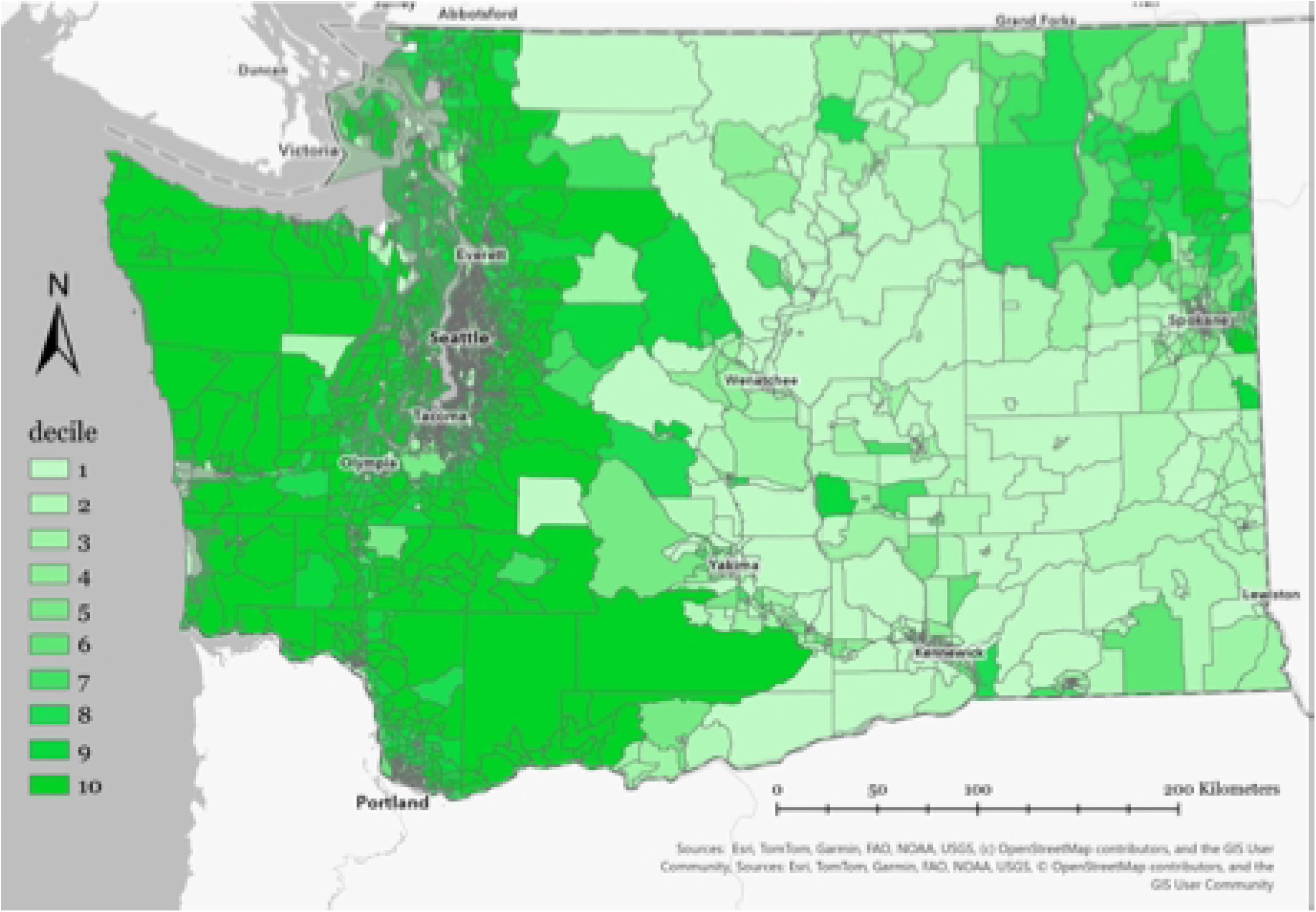
2020 Washington NDVI distribution by deciles Note. Block groups are shaded by population-weighted NDVI decile (Decile 1 = least green; Decile 10 = greenest).

This skewed distributional characteristic has clear methodological consequences for the benefit transfer model. Because Washington’s population is concentrated in the higher greenness deciles, the normalization technique described in Equations (1)-(3) yields a statewide cost index anchored to a higher baseline greenness. The normalizing constant *α* adjusts downward to account for Washington’s population-weighted average NDVI decile exceeding the reference category (Decile 1). This reduces the range of absolute cost savings associated with incremental greening compared to a less-vegetated population. Consequently, the healthcare cost savings reported in this study should be interpreted as conservative lower-bound estimates; the true population-level savings attributable to greenspace access in Washington are likely larger than those reported by the current benefit transfer framework.

The estimated population-weighted per-capita healthcare costs for insured Washington residents in 2020, by residence NDVI decile, show a broadly inverse relationship between residential greenness and per-capita healthcare expenditure (Table 1, Fig 2). This finding is consistent with the dose-response gradient documented in the source study by Van Den Eeden et al. [33] and supported by a larger body of epidemiological evidence linking residential vegetation to lower morbidity burden and healthcare utilization [17,18]. Residents in the least-green decile (Decile 1) have an estimated average annual healthcare cost of $6,472.75 per capita, while those in the greenest residential category (Decile 10) have an average of $5,939.39, a $533.35 difference per person per year (Table 1). The gradient is nearly monotonic across the decile range, with per-capita expenditures falling gradually from Decile 1 to Decile 10. One notable exception is the shift from Decile 3 ($6,378.24) to Decile 4 ($6,390.54), where estimated costs marginally increase by approximately $12.30 per capita. This localized non-monotonicity is small in absolute magnitude and may reflect residual heterogeneity in population composition or the spatial distribution of sociodemographic risk factors, such as concentrated poverty, elevated chronic disease burden, or limited access to preventive care, within mid-range NDVI block groups. Critically, this local reversal does not deviate from the overall downward trend, and the ten-decile pattern robustly supports the conclusion that higher residential greenness is systematically associated with lower per-capita healthcare spending across Washington.

**Fig 2.**
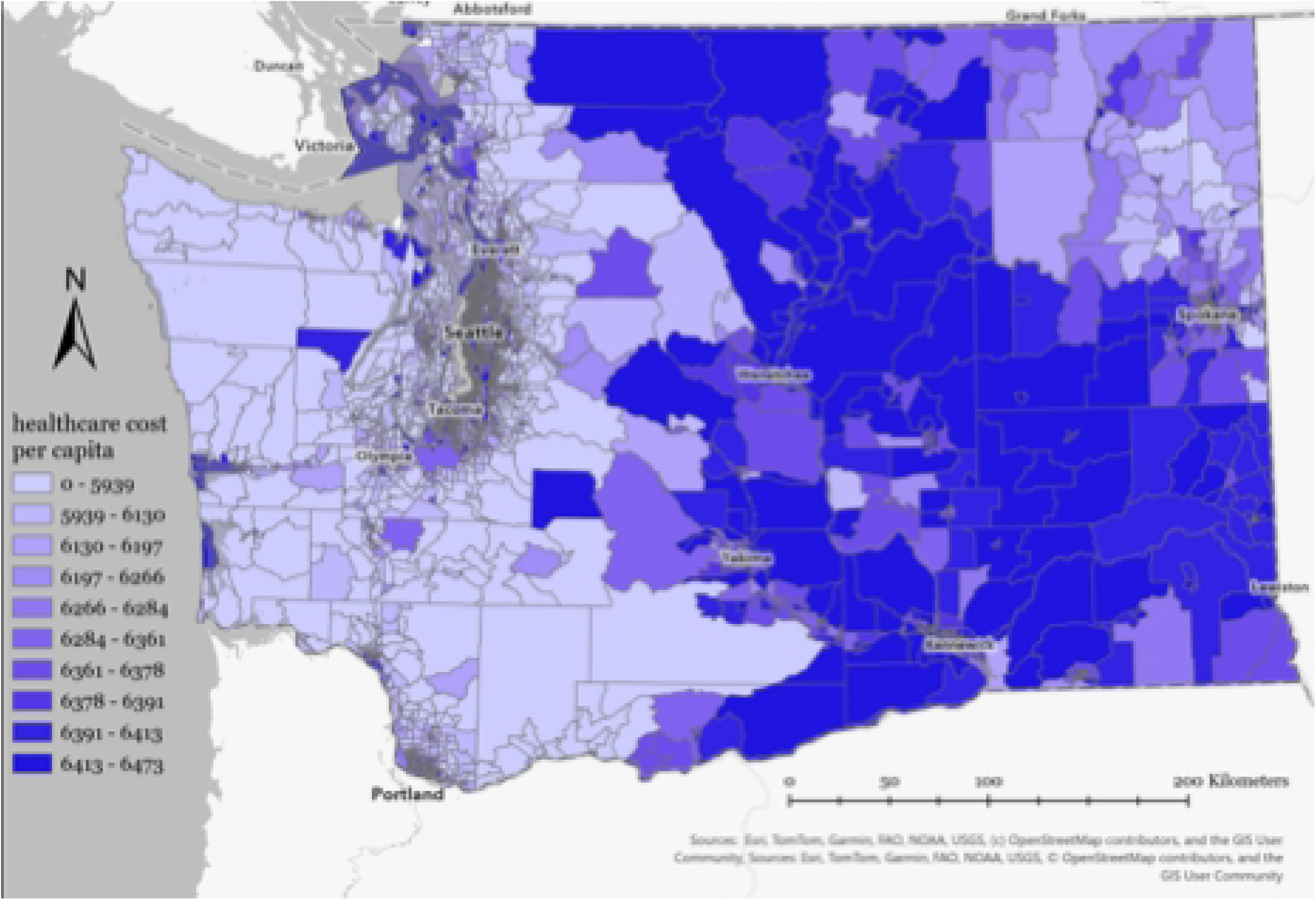
2020 Washington estimated healthcare cost per capita by decile

**Table 1.** Washington Population-Weighted Per-capita Healthcare Cost.

| Decile | Average Per-capita Healthcare Cost | 95% CI Per-capita Healthcare Cost | Population | Minimum NDVI | Median NDVI | Maximum NDVI |
| --- | --- | --- | --- | --- | --- | --- |
| 1 | \$6,472.75 | \$6,443.24–\$6,502.59 | 758,327 | 0.021 | 0.270 | 0.330 |
| 2 | \$6,412.55 | \$6,325.77–\$6,499.72 | 734,754 | 0.330 | 0.372 | 0.400 |
| 3 | \$6,378.24 | \$6,293.71–\$6,465.91 | 769,007 | 0.400 | 0.425 | 0.451 |
| 4 | \$6,390.54 | \$6,298.13–\$6,482.73 | 716,895 | 0.451 | 0.470 | 0.488 |
| 5 | \$6,360.77 | \$6,271.29–\$6,451.49 | 747,434 | 0.488 | 0.505 | 0.523 |
| 6 | \$6,283.74 | \$6,188.61–\$6,376.65 | 740,841 | 0.523 | 0.541 | 0.558 |
| 7 | \$6,265.62 | \$6,171.30–\$6,357.78 | 776,251 | 0.558 | 0.575 | 0.595 |
| 8 | \$6,197.01 | \$6,109.68–\$6,287.14 | 875,815 | 0.595 | 0.620 | 0.648 |
| 9 | \$6,130.34 | \$6,042.39–\$6,219.35 | 832,386 | 0.648 | 0.679 | 0.710 |
| 10 | \$5,939.39 | \$5,851.29–\$6,029.09 | 739,167 | 0.711 | 0.755 | 0.878 |

*Note. CI stands for confidence interval. NDVI stands for normalized difference vegetation index. Costs are annual 2020 U.S. dollars.*

Examining the cost gradient throughout the whole distribution reveals a key structural feature: the marginal savings per increasing greenness decile are not uniform. The per-capita cost decrease from Decile 1 to Decile 5, which covers the bottom half of the greenness distribution, is roughly $111.98 per person per year, whereas the reduction from Decile 5 to Decile 10 is approximately $421.38 per person per year. This nonlinear dose-response pattern suggests that as residential vegetation increases, healthcare cost advantages may accelerate significantly, especially above approximately Decile 5. This pattern is consistent with the underlying epidemiology of conditions most responsive to sustained greenspace contact, such as anxiety, depression, and cardiovascular disease, where more pronounced reductions in healthcare utilization tend to occur at moderate-to-high nature doses rather than at lower thresholds [1,4].

The statewide per-capita cost difference of $533.35 between Decile 1 and Decile 10 in Washington is larger than Van Den Eeden et al.’s [33] estimate of $374.04. This discrepancy does not reflect a stronger greenness-health association in Washington; rather, it is a mathematical consequence of the state’s higher calibrated baseline healthcare expenditure. Because the transferred rate ratios are dimensionless multipliers of a baseline cost level (Equation 2–3), the same relative gradient produces a proportionally larger absolute dollar spread wherever that baseline is higher. Washington’s calibrated statewide average per-capita cost $6,280.17 [58] exceeds the baseline implied by the Kaiser Permanente source population, and this difference in baseline cost level, not a difference in the strength of the underlying dose-response relationship, accounts for the larger absolute gap observed here. The transferred RRs are identical across both regions by design; this is the foundation of the benefit transfer logic. However, their conversion into absolute dollar differentials is governed entirely by Washington’s own per-capita healthcare expenditure baseline, not by any feature of the state’s greenness distribution. Only the decile-specific rate ratios are carried over from the source study, while the regression intercept, Northern California’s baseline cost levels, covariate–cost relationships, and health-system utilization patterns are explicitly excluded. The dollar figures reported here therefore reflect Washington’s own cost structure rather than an imported one.

This result has important implications. The observed disparity of $533.35 represents marginal healthcare savings associated with moving from the least green to the most green decile within Washington’s existing distribution of greenspace, rather than the total health benefits that greenspace provides relative to a hypothetical unvegetated environment [59]. Because Washington already contains an network of parks, forests, and riparian corridors with extensive tree canopies, these existing natural assets are likely already generating substantial health benefits for residents [60]. Consequently, the decile-differential metric captures only the additional savings attributable to variation within the current greenness distribution, while the baseline health benefits provided by Washington’s existing green infrastructure remain unobserved and therefore are not included in this analysis. Future research that uses longitudinal data to link changes in NDVI to individual-level claims data over time could better isolate the dynamic contribution of greenspace change to healthcare cost trajectories across the state, potentially revealing a richer and larger total benefit than the cross-sectional cost gradient reported here.

Benchmarked against other economic estimates of greenspace’s healthcare value, our findings fall within a plausible range. Becker et al. [32], using county-level forest cover and Medicare enrollees, found that each one-percentage-point increase in forest cover was associated with $4.32 less in per-capita Medicare spending, extrapolating to $6–9 billion in national annual savings; their coarser exposure measured an older, fee-for-service population versus the individual-level, all-payer population used here. Kondo et al. [34] recently evaluated associations between mean NDVI at the census tract level and healthcare costs in a longitudinal study of Medicare expenditures among fee-for-service clients (ages 65+; n = 275,000) across the US. They found average annual claim totals were $957.30 lower for participants living in the most green compared to the least green areas. Feng et al. [30] followed 110,134 Australian adults longitudinally and estimated that raising tree canopy from below 10% to at least 30% would save AU$193 per person annually in cardiovascular-related hospital costs alone (AU$569 among those hospitalized); because this captured only cardiovascular hospitalizations, it represented a lower bound for a single condition rather than a benchmark for our all-cause cost gradient. Loch et al. [61] combined mobile-device visitation data with self-reported health differences to estimate that access to biodiverse greenspace in metropolitan Adelaide generated approximately AU$140 million in reduced healthcare costs in 2018–2019. This was about 4% of the region’s chronic-disease healthcare budget, a share larger than the 0.1–0.3% of statewide expenditure implied by our scenario-based estimates ($45.6–$157.7 million) although our estimated savings per affected resident are approximately 0.8–1.0% of the statewide per-capita baseline of $6,280. Despite differing countries, health systems, exposure measures, and outcomes, these studies converge on a common conclusion: greenspace is associated with meaningful but modest reductions in healthcare spending, typically a few percent of total costs, placing our Washington estimates well within ranges from this emerging literature.

Table 2 compares three hypothetical greening scenarios, each targeting a different segment of Washington’s NDVI distribution, and together they illustrate a familiar tension in spatially targeted public health interventions: the trade-off between reach and intensity. The baseline annual mean per-capita healthcare cost for all scenarios is $6,280.17. Scenario 1 (Bottom-Decile Lift), which reassigns only the 477 CBGs in the lowest greenness decile, produces the smallest total effect ($45.6 million annually) but the largest per-resident benefit among the affected population ($60.20 per person per year). Scenario 2 (Lower-Quintile Uplift), which extends the same style of intervention across the bottom four deciles, or roughly 40% of the CBGs in Washington, generates more than three times the total statewide savings ($157.7 million annually), but at a lower average benefit per affected resident ($52.95). This pattern reflects the different counterfactual each scenario applies, not a difference in the steepness of the cost gradient itself: Scenario 1 lifts Decile 1 only as far as Decile 2, capturing a $60.20 per-capita gap (Table 1), whereas Scenario 2 lifts Deciles 1 through 4 all the way to Decile 5, a larger jump for Decile 1 residents ($111.98) but a much smaller one for Deciles 2 through 4, whose costs already sit close to the Decile 5 level ($51.78, $17.47, and $29.77, respectively). Averaging across this heterogeneous group pulls Scenario 2’s per-resident benefit below Scenario 1’s, even as its broader reach more than triples the total statewide savings.

**Table 2.**
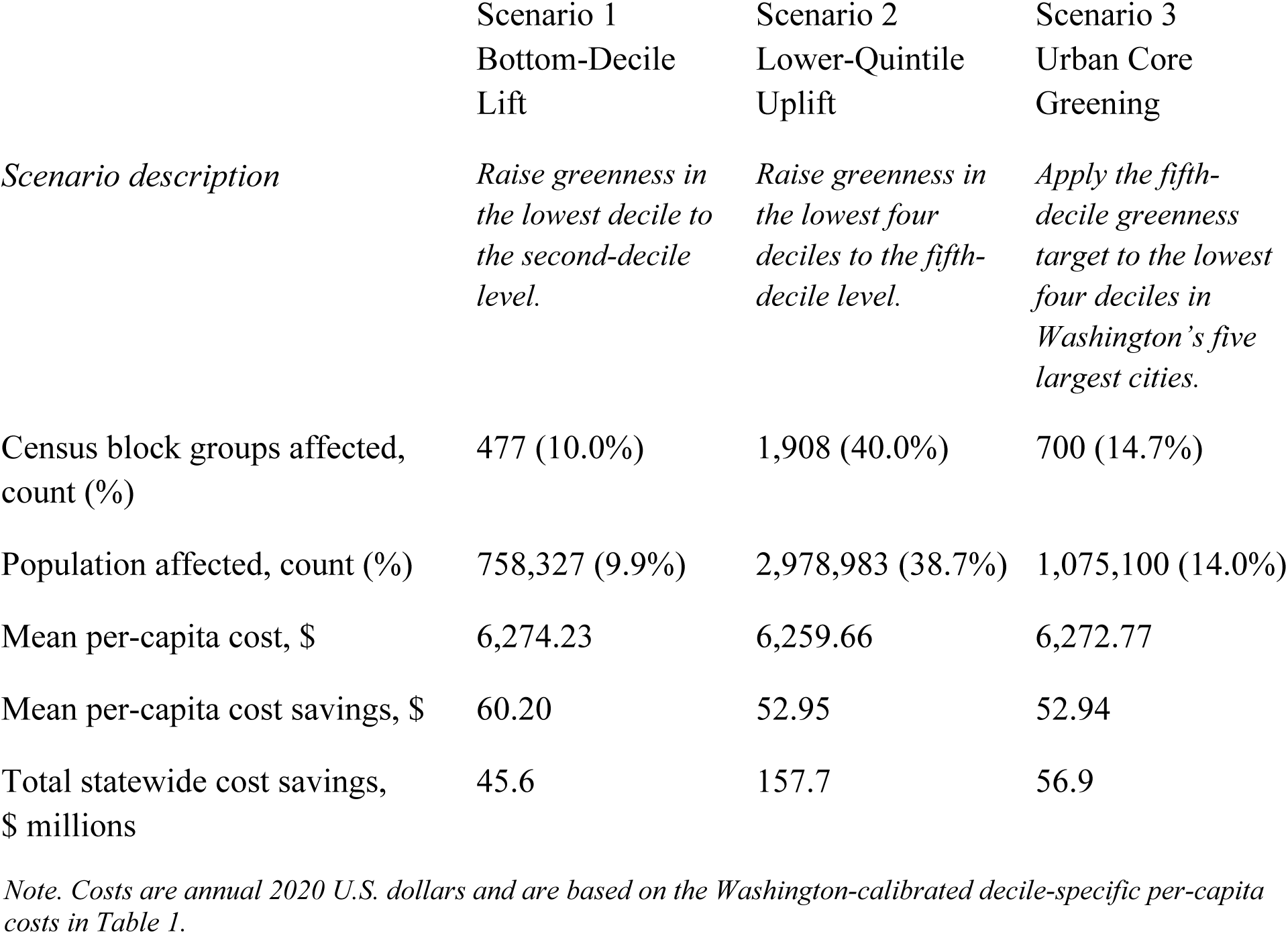
Washington Healthcare Cost Greening Scenario Simulations.

|  | Scenario 1<br>Bottom-Decile<br>Lift | Scenario 2<br>Lower-Quintile<br>Uplift | Scenario 3<br>Urban Core<br>Greening |
| --- | --- | --- | --- |
| *Scenario description* | *Raise greenness in the lowest decile to the second-decile level.* | *Raise greenness in the lowest four deciles to the fifth-decile level.* | *Apply the fifth-decile greenness target to the lowest four deciles in Washington's five largest cities.* |
| Census block groups affected, count (%) | 477 (10.0%) | 1,908 (40.0%) | 700 (14.7%) |
| Population affected, count (%) | 758,327 (9.9%) | 2,978,983 (38.7%) | 1,075,100 (14.0%) |
| Mean per-capita cost, \$ | 6,274.23 | 6,259.66 | 6,272.77 |
| Mean per-capita cost savings, \$ | 60.20 | 52.95 | 52.94 |
| Total statewide cost savings, \$ millions | 45.6 | 157.7 | 56.9 |

*Note. Costs are annual 2020 U.S. dollars and are based on the Washington-calibrated decile-specific per-capita* *costs in Table 1.*

Scenario 3 (Urban Core Greening), which applies the identical Decile-5 uplift rule as Scenario 2, but restricts it to CBGs within Washington’s five largest cities, offers a useful internal check on the model: its mean savings per affected resident ($52.94) are nearly indistinguishable from Scenario 2’s statewide average ($52.95), despite drawing on a geographically narrower and demographically distinct subset of block groups. This similarity indicates that the marginal cost-per-decile relationship estimated from the full state sample generalizes well to the urban subpopulation, and that the urban cores are not systematically composed of outlier deciles relative to the state as a whole. In absolute terms, Scenario 3 reaches roughly 1.08 million residents, representing 14.0% of the state’s population, a larger relative share than Scenario 1’s 9.9%. Consistent with this broader geographic footprint, Scenario 3 also produces a larger total statewide benefit ($56.9 million) than Scenario 1, though the difference between the two scenarios remains modest relative to the gap in population coverage.

Several limitations are relevant to our research. First, benefit transfer is, by construction, an exercise in informed approximation rather than direct measurement: we assumed that the relationship between green cover and healthcare cost estimated among Kaiser Permanente members in Northern California holds with the same functional form in Washington, an assumption we defended on ecological and institutional grounds without Washington-specific claims data. Second, our analysis is cross-sectional, built on a single year of 2020 cost and exposure data; it can describe a static gradient but cannot speak to how healthcare costs respond as a given block group’s greenness changes over time, nor can it fully disentangle greenness from the sociodemographic and infrastructural factors that often cluster alongside it. A related limitation concerns temporal alignment: our Washington data are from 2020, the most recent available, while the source rate ratios were estimated from Kaiser Permanente Northern California costs spanning 2003–2015 [33] and are applied here unadjusted for that gap. We therefore cannot rule out that the underlying greenness–cost relationship has shifted since the source data were collected, due to changes in healthcare delivery, chronic disease prevalence, or the built environment. Third, because the source study’s rate ratios were estimated relative to Northern California’s least-green decile, and Washington is greener than that reference population, our decile-differential estimates likely understate the total economic value of the state’s greenspace, particularly at the high end of the distribution where our data cannot extrapolate. Finally, while the source study found no significant interaction between greenness and sociodemographic covariates, we were not able to test whether this homogeneity holds within Washington, and our statewide and scenario-level estimates therefore say little about how these savings, or the burden of their absence, are distributed across the state’s most vulnerable communities.

Each of these limitations point toward a concrete next step. The most valuable is not simply a longitudinal test, but one that improves on a recent attempt at exactly this design: Browning et al. [62] linked 13 years of Kaiser Permanente Northern California claims to residential moves and found no significant associations, a null result the authors attribute to self-selected relocation, a follow-up period likely too short to capture chronic-disease latency, and confounding between movers and non-movers. The most valuable next step is therefore a Washington-specific study linking NDVI change from an exogenous source, such as a municipal tree-planting or park-development program, to individual-level medical claims over a longer follow-up period, isolating the dynamic contribution of greening to healthcare costs without these limitations, and potentially revealing a larger total benefit than the cross-sectional gradient captured here. A second priority is an explicitly equity-focused extension of this framework, stratifying the cost-savings estimates by race, income, and historical redlining status to determine whether the health dividends of greening, and the risks of green gentrification that can accompany it, are distributed fairly across the communities that stand to benefit most. A third is mechanistic: pairing economic estimates like ours with the emerging biomedical literature on immune, microbiome, and neuroendocrine pathways could help clarify why the dose-response curve we found accelerates above the state’s median greenness, and could sharpen predictions about which interventions yield the largest health returns per dollar invested. Finally, because the benefit transfer framework introduced here relies only on a transferable dose-response relationship and each state’s own exposure and cost data, it offers a template that other states, and eventually national entities, could adopt to build a fuller, comparable picture of greenspace’s economic contribution to public health across the country.

## Conclusion

This study set out to answer a question that has long lingered at the edge of ecosystem-service research: what is greenspace worth to healthcare systems, expressed in dollars that a legislator, health plan, or city budget office can use to inform decision making? By transferring the decile-specific cost ratios estimated among nearly five million Kaiser Permanente members in Northern California onto Washington’s population-weighted greenness distribution, we found that residents in the state’s least-green census block groups incur roughly $533 more in annual per-capita healthcare costs than residents in the greenest block groups, on average. This $533 cost gradient does not increase linearly with greenness. The marginal savings from becoming greener accelerate as greenness increases, with the upper half of Washington’s greenness distribution delivering nearly four times the cost reduction of the lower half. Translating this pattern into policy-relevant greening scenarios, it produced avoided healthcare expenditures ranging from $45.6 million a year for a narrowly targeted lift of the state’s least-green block groups to $157.7 million a year for a broader uplift spanning the bottom two-fifths of the distribution, with a comparably sized urban-core intervention yielding benefits nearly indistinguishable, per resident, from the statewide average. Reallocating scarce greening investment, in other words, is not only an ecological or aesthetic choice; it is a decision with a measurable economic benefit attached to it. These healthcare cost reductions likely represent only a portion of the total value greening investments generate, since benefits such as stormwater management, urban heat mitigation, carbon sequestration, and property value gains fall outside the scope of this analysis. At the same time, because this study estimates only the benefit side of the ledger, it cannot speak to whether any particular greening intervention would pass a full benefit-cost test; the answer depends on implementation costs that vary substantially by location, land availability, and intervention type, and that we do not model here.

Public infrastructure budgets involve competing demands for funding of sidewalks, sewer lines, and school renovations, as well as green infrastructure. The latter can best be integrated into spending, investment, and policy decisions when its returns can be stated in the same units as its costs. This study contributes such evidence by applying an observed dose-dependent relationship between greenness and healthcare costs to estimate potential avoided spending in Washington. We also note the importance of avoiding overreach: because Washington’s population already sits in a greener part of the exposure spectrum than the Northern California sample from which our rate ratios were drawn, our estimates should be read as a conservative floor rather than a ceiling. The true value of the state’s existing tree canopy, parks, and riparian corridors, most of which lie above the greenness range observed in the original study, likely exceeds what a decile-to-decile comparison alone can reveal.

Urbanization and a changing climate are highlighting, and in some cases exacerbating, the association between where people live and the environmental stressors that erode their health, even as the evidence for nature’s restorative power continues to grow. What is often missing in policy contexts is not proof that greenspace matters insofar as it improves physical and mental health, but a way of demonstrating how much it matters in terms in which public budgets already operate. This study offers Washington one relevant set of findings, along with a method for generating others. It is not intended as a final or definitive number, but as a transparent, defensible starting point for measuring the health-related value of trees, parks, and riparian corridors for individuals who live near them.

## Data and code Availability

Code used for this project is available from the corresponding author upon reasonable request and will be deposited in a public repository upon acceptance.

## Data Availability

Data and code used for this project is available from the corresponding author upon reasonable request and will be deposited in a public repository upon acceptance.

## Acknowledgements

This work is supported by the McIntire-Stennis (MS) Cooperative Forestry Research Program from the U.S. Department of Agriculture’s National Institute of Food and Agriculture under accession number 7010196. We thank Stephen K. Van Den Eeden for clarifying the methods and providing decile-specific rate ratios of the source study, and Leo Wahl for assistance with NDVI data preparation.

## Ethics statement

This study used only publicly available, aggregated data and did not involve human participants or identifiable personal information; institutional review board approval was therefore not required.

## Statement on use of artificial intelligence

During the preparation of this manuscript, the authors used OpenAI’s GPT-4 to assist with language editing, including grammar checking, formatting, and copyediting, as well as to identify potential errors in computer code. The authors independently reviewed and verified all AI-assisted outputs and remain fully responsible for the content, accuracy, interpretation, and integrity of the manuscript.

